# Subsumption, Vectorization, Heat Maps, and Word Clouds Support the Visualization of Orphadata Neurology Phenotypes

**DOI:** 10.1101/2022.12.22.22283847

**Authors:** Daniel B. Hier, Raghu Yelugam, Michael D. Carrithers, Donald C. Wunsch

**Author notes:** Correspondence: Daniel B. Hier.

## Abstract

Disease phenotypes are characterized by signs (what a physician observes during the examination of a patient) and symptoms (the complaints of a patient to a physician). Large repositories of disease phenotypes are accessible through the Online Mendelian Inheritance of Man, Human Phenotype Ontology, and Orphadata initiatives. Many of the diseases in these datasets are neurologic. For each repository, the phenotype of a neurologic disease is represented as a variable-length list of concepts selected from a suitable ontology. Visualizations of these lists are not provided. We address this limitation by using subsumption to collapse the number of descriptive features from 2,946 classes into thirty superclasses. Phenotype feature lists of variable lengths were converted into fixed-length numerical vectors. Phenotype vectors can be aggregated into matrices and visualized as heat maps that allow side-by-side disease comparisons. Individual diseases (representing a row in the matrix) can be visualized as word clouds. We illustrate the utility of this approach with a use case based on 32 dystonic diseases in Orphadata. The use of subsumption to collapse phenotype features into superclasses, the conversion of phenotype lists into vectors, and the visualization of phenotypes vectors as heat maps and word clouds contribute to the improved visualization of neurology phenotypes in Orphadata.

## INTRODUCTION

The signs and symptoms of a disease characterize its phenotype. In addition to signs (what a physician observes in a patient) and symptoms (the complaints of a patient), a clinical phenotype can include the age of onset of a disease, its mode of onset, its rate of progression, its mode of inheritance, and its response to treatment. Some researchers include biochemical, radiological, electrophysiological, and biosensor findings as part of the disease phenotype [26, 22, 11, 4, 36]. Large phenotype registries are available on the internet. The On-Line Mendelian Inheritance in Man (OMIM) repository has over 9,500 disease profiles [2] and Orphadata has phenotype profiles of 4,245 rare diseases [33]. The Human Phenotype Ontology (HPO) draws phenotype profiles from Orphadata and OMIM so that some genetic diseases have alternative profiles from each registry [20, 29]. All three repositories have sophisticated search engines that retrieve phenotype features by disease or gene [26]. Phenotypic features are recorded as concepts (terms) from restricted vocabularies such as the Human Phenotype Ontology (20,246 terms) [31], or the Online Mendelian Inheritance of Man ontology (99,165 terms) [34].

### Neuro-phenotypes

Deep phenotyping is the detailed description of the signs and symptoms of a patient or a disease utilizing a restricted vocabulary [36]. Deep phenotypes are represented as linear lists of signs and symptoms and their associated machine codes from an ontology such as HPO or OMIM (see, for example, Table 1). The June 2022 release of Orphadata lists 7,261 rare diseases and classifies 1,740 as rare neurological diseases (https://www.orphadata.com/linearisation/). Orphadata provides phenotype profiles on 4,254 rare diseases, of which 1,184 are neurologic (https://www.orphadata.com/phenotypes/). Neuro-phenotyping is the deep phenotyping of neurological disease [23]. We have previously suggested that most neuro-phenotyping can be done with a restricted vocabulary of about 1,600 concepts [24].

**Table 1.**
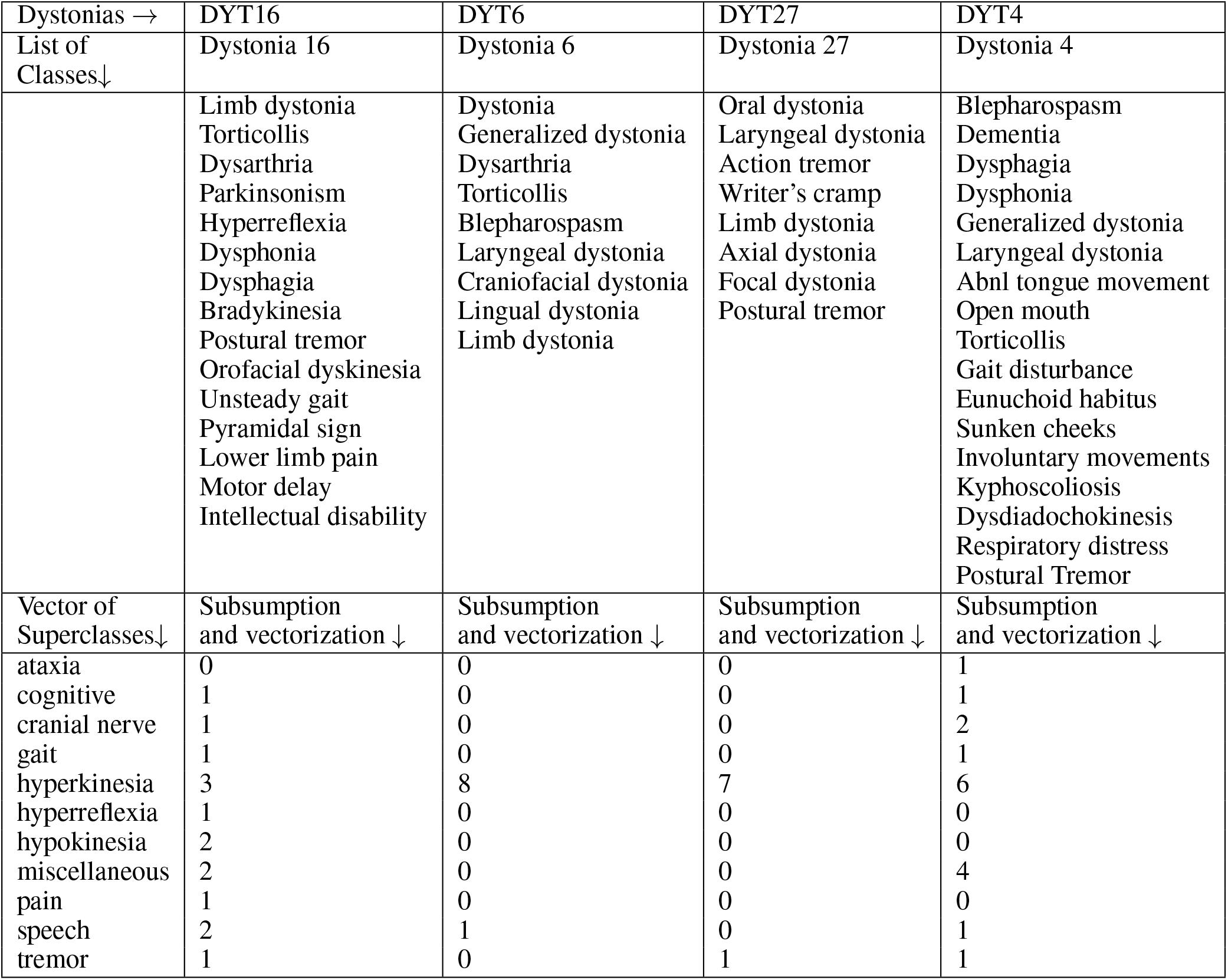
The upper half of the Table shows lists of signs and symptoms for each dystonic disease from Orphadata. In the lower half of the Table, lists of classes have been converted to vectors of superclasses using subsumption governed by a lookup table. Counts are the number of times each class occurs in the superclass and is the input for the row values for the heat maps. Columns from the top half are variable length lists; columns from the bottom half are fixed length vectors.

Although lists of phenotypic features for neurological diagnoses can be retrieved from Orphadata, OMIM, or HPO, these lists are difficult to visualize. For example, the Orphadata annotations for Dystonia Type 13 (DYT13) are:

*Very frequent*

Stereotypy

Torsion dystonia

Torticollis

*Frequent*

Limb dystonia

Dystonia

Craniofacial dystonia

Jerky head movements

*Occasional*

Postural tremor

Action tremor

Focal dystonia

*Rare*

Generalized dystonia

Hoarse voice

These lists have limitations. The lists may be long. In the Orphanet dataset, 25% of the lists are more than 34 features. These lengths are beyond the length of 7*±*2 that is easily comprehended [35]. Side-by-side comparisons of these lists are difficult (Table 1). Lists of signs and symptoms from Orphadata may contain pathologies (e.g., gliosis, Lewy bodies), radiological findings (e.g., abnormal PET FDG), biochemical findings, electrophysiological findings, and modes of inheritance. Although terms in Orphadata are from the HPO-controlled vocabulary (20,246 classes) [31], redundancies, near-synonyms, hypernyms, and hyponyms populate the lists (e.g., dysarthria and slow slurred speech; bradykinesia and hypokinesia, masked facies and hypomimia, etc.) Furthermore, OMIM, Orphadata, and HPO do not provide native methods for visualization of phenotype lists.

### Prior work

Limited work has been done on visualizing phenotype lists retrieved from HPO, OMIM, or Orphadata. Xu et al. [40] visualized the distances between genetic diseases and their underlying phenotypes using t-SNE (stochastic neighborhood embedding) maps. The phenotype features from the OMIM dataset were used to calculate distances between genetic diseases. The t-SNE maps are a 2-dimensional representation of the distances between genetic diseases derived from multi-dimensional data. Although these t-SNE maps provide instructive information about the distances between genetic diseases, they do not reveal the details of the underlying phenotypes. Network analysis and network graphs have been used to visualize the distances between diseases based on their phenotype [14, 37, 30]. However, these network diagrams do not elucidate the underlying phenotypic differences between the diseases. Several methods have been proposed to visualize disease-phenotype relationships, including radar graphs [10], co-occurrence charts [16], and sunburst diagrams [17]. Cao et al. have developed visualization techniques called DICON, FacetAtlas, and SolarMap that show promise for visualizing phenotype features by disease [7, 5, 6, 18].

An additional barrier to visualizing neurology phenotype profiles is the large number of terms in the HPO (N=20,390), making the number of columns in heat maps or tables that visualize phenotypes impractical. A feature reduction strategy that chunks phenotype features into a more manageable number of superclasses is needed. For example, Hier and Pearson [25] have suggested chunking problems in the electronic health record by body system to increase the readability of the problem list. Both OMIM and HPO chunk phenotype features by body system. Orphanet chunks phenotype features by feature frequency (common to rare). Yauy et al. [41] have chunked 16,600 phenotypic traits into 390 interacting symptom groups. However, the chunking of phenotype features by body system is unlikely to yield useful visualizations because dissimilar phenotypic features are grouped. For example, chunking concepts by nervous system category would put the unlike concepts of hypertonia, hypotonia, hyperreflexia, and hyporeflexia into the same category, a grouping of little diagnostic value. Although the chunking of phenotype concepts by body system or other schemes helps organize phenotype features, it does not reduce the number of features. Since the HPO is a hierarchical containment ontology, we have suggested that subsumption can create superclasses of phenotypic features and reduce the number of features [39, 38].

### Proposed Approach and Use Case

We propose to improve the visualization of neurology phenotypes in the Orphdata dataset utilizing a combination of subsumption, vectorization, heat maps, and word clouds. As proof of concept, we illustrate the utility of this approach with a use case that visualizes the phenotype lists of 32 dystonic diseases from Orphadata. In 1911 Oppenheim described the disease *dystonia musculorum deformans* and coined the term dystonia [21]. Albanese et al. [1] defined dystonia as “a rare movement disorder characterized by sustained or intermittent muscle contractions causing abnormal, often repetitive movements, postures, or both.” Since the description of dystonia by Oppenheim, many forms of dystonia have been described. Dystonia is classified along two axes: clinical and etiologic [1]. Clinical classification is by age at onset, body distribution, the temporal pattern of symptoms, and associated phenotype features. Etiologic classification is by genetic versus non-genetic causation. Dystonia is one of the hyperkinetic movement disorders which also encompasses chorea, athetosis, hemiballismus, tics, tremors, stereotypy, myoclonus, and dyskinesia [28]. Although diseases labeled a *dystonia* have a core symptom of dystonia, there is considerable variability in the clinical presentation (signs and symptoms) of the dystonias [21, 32, 13], making it an excellent use case for phenotype visualization. Furthermore, better characterization and classification of the dystonias is a major initiative of the European Reference Network for Rare Diseases, and Orphadata [8, 19].

We downloaded the most recent Orphadata file with phenotype annotations of rare diseases, including those classified as rare neurological diseases. We identified the unique HPO terms used to characterize the signs and symptoms of rare neurological diseases and created a lookup table to map each term to one of 30 superclasses based on subsumption and expert opinion. The lists of phenotypic features for 32 dystonic diseases from Orphadata were converted into phenotype vectors, with the first element of the vector being the disease name and the next 30 elements being the count of features (signs and symptoms) for each superclass. The matrix of phenotype vectors can be visualized as a heat map (Figure 1); individual rows (vectors) can be visualized as word clouds (Figure 2b).

**Figure 1.**
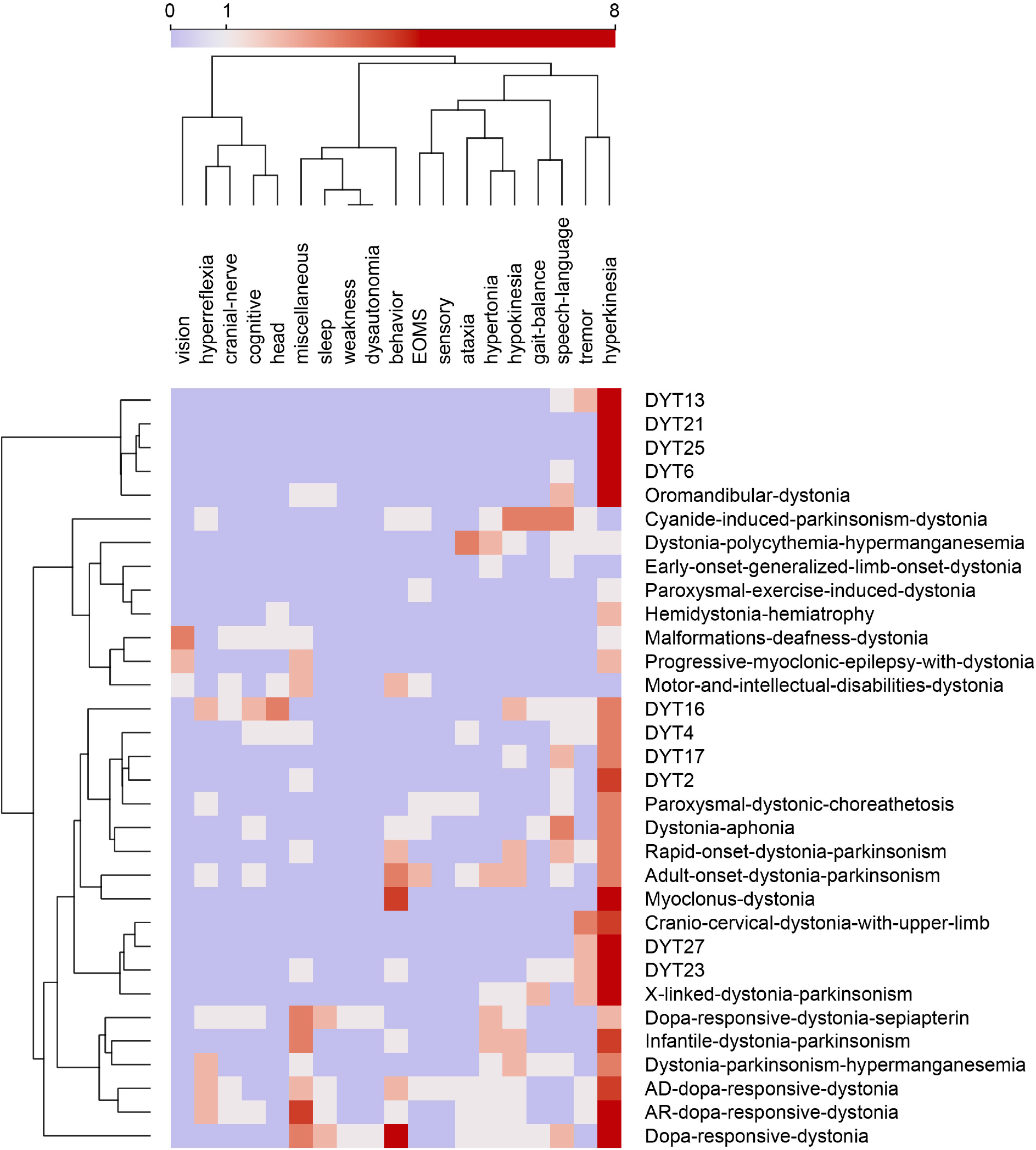
Heat map of 32 dystonias from Orphadata. Each row is a different dystonic disease. Each column is one of 19 phenotype superclasses. Counts in columns range from 0 to 8. The color scale is centered at 1. Rows and columns are clustered by hierarchical clustering with Ward linkage. Distances between columns are by Pearson correlation coefficient. Distances between rows are by Euclidean distance. Hyperkinesia is the largest superclass, followed by tremor, behavior, hypokinesia, speech_language, and miscellaneous (See word cloud in Figure 3a).

**Figure 2a.**
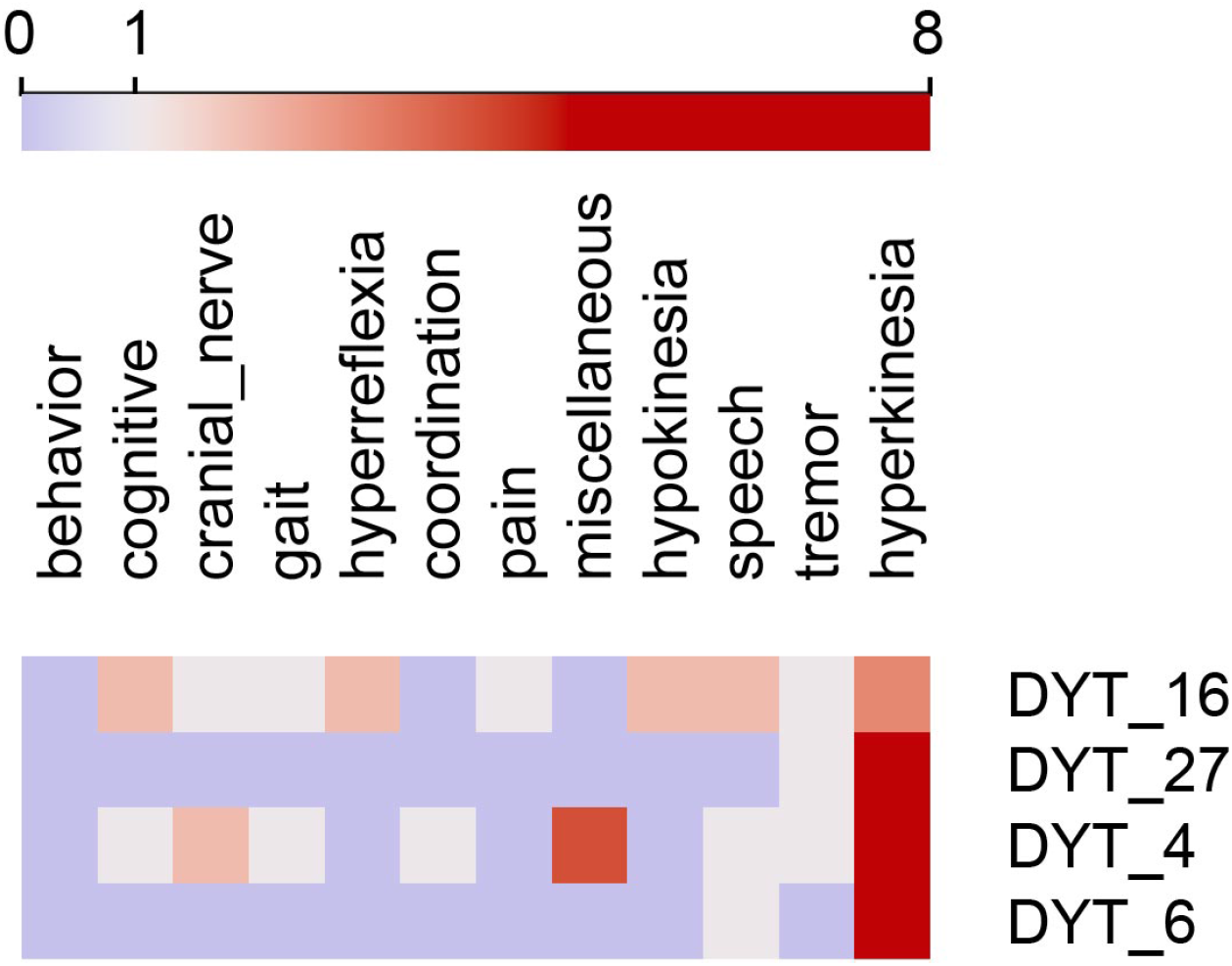
Heat map of four selected cases of dystonia. Columns are feature superclasses, and rows are diseases. Heat maps and word clouds are based on Table 1. Each row in the heat map represents a column of signs and symptoms from Table 1. Feature scores range from 0 to 8, with the color scale centered at 1. There is no row or column clustering. Word cloud visualizations of each row are below.

**Figure 2b.**
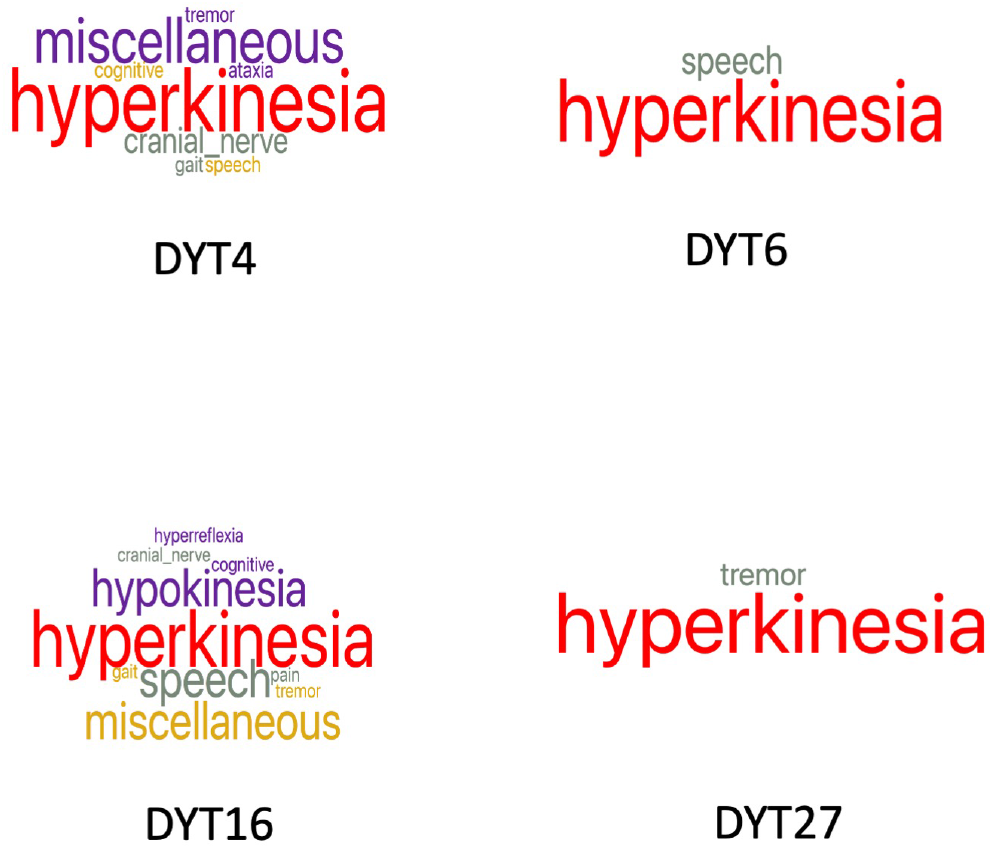
Word clouds for the four forms of dystonia represented in the heat maps above. Word size reflects the feature count in each superclass. DYT6 and DYT27 are pure dystonia, whereas DYT4 and DYT16 have other non-dystonic features.

**Figure 3a.**
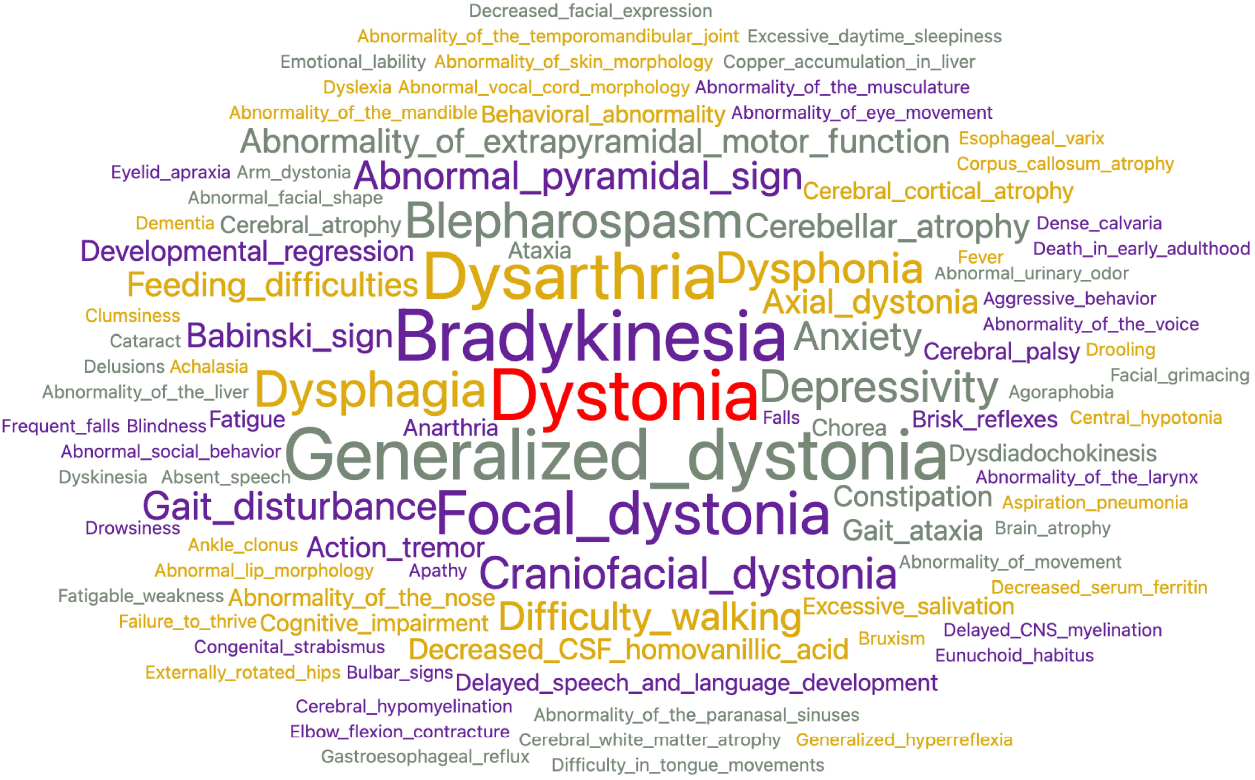
To characterize the 32 dystonic diseases, 528 total concepts and 252 unique concepts were used. The most frequent concepts used were dystonia, bradykinesia, generalized dystonia, dysarthria, and focal dystonia.

## METHODS

### Data acquisition (Phenotype feature lists)

The XML file with 4254 rare disease disorders and 112,256 phenotypic annotations was downloaded (June 2022 release of Orphadata: (*https://www.orphadata.com/phenotypes/*). Phenotype features are coded using the HPO ontology. Orphadata defines a rare disease as affecting less than 1 in 2,000 individuals in Europe and classifies 1184 of the diseases as rare genetic neurological diseases. We used python to parse the XML file and create a variable-length list of phenotypic features for each disease. We retained phenotypic annotations that were clinical signs or symptoms and filtered out phenotypic annotations related to disease course (progressive, static, etc.), mode of inheritance (recessive, dominant, etc.), biochemical abnormality, radiological abnormality, pathological abnormality, or electrophysiological abnormality. Based on published literature, Orphadata classifies the frequency of each phenotypic feature from rare (1 to 4%) to always present (100 %). We retained phenotypic features classified as occasional or higher (5 to 100 %).

### Subsumption (Conversion of phenotype classes to superclasses)

The HPO [31] is organized as a hierarchical subsumption ontology so that more-specific concepts in the ontology are subsumed by more general concepts [38]. We identified 2,946 unique concepts that Orphadata used to phenotype neurological diseases. We collapsed these concepts into 30 superclasses using subsumption and domain expert opinion. Example class memberships (three classes for each superclass) and superclass counts are shown below.

1. alertness (53 terms) delirium, drowsy, somnolence
2. ataxia (62 terms) asynergia, clumsiness, dystaxia
3. atrophy (69 terms) muscle atrophy, atrophy, limb fasciculations
4. behavior (238 terms) apathy, anxiety, delusions
5. cognitive (202 terms) agnosia, apraxia, forgetfulness
6. cranial nerve (203 terms) ageusia, hyperacusis, facial diplegia
7. dysautonomia 35 terms) hypohidrosis, orthostatic syncope, dysautonomia
8. eye movements (272 terms) upgaze palsy, nystagmus, hypometric saccades
9. fatigue (26 terms) muscle fatigue, fatigable weakness, fatigue
10. gait (110 terms) ataxic gait, falls, unsteady gait
11. head (263 terms) microcephaly, macrocephaly, increased head size
12. hyperkinesia (157 terms) dyskinesia, dystonia, hyperkinesia
13. hyperreflexia (58 terms) increased reflexes, clonus, hyperreflexia
14. hypertonia (58 terms) increased muscle tone, rigidity, spasticity
15. hypokinesia (66 terms) bradykinesia, akinesia, hypomimia
16. hyporeflexia (43 terms) areflexia, hyporeflexia, absent ankle reflex
17. hypotonia (19 terms) decreased tone, muscle flaccidity, limb hypotonia
18. other muscle (119 terms) myokymia, muscle hypertrophy, myotonia
19. neck (48 terms) stiff neck, neck rigidity, meningismus
20. pain (145 terms) pain, arm pain, allodynia
21. seizure (358 terms) seizure, tonic-clonic seizure, febrile seizure
22. sensory (192 terms) hyperesthesia, dysesthesia, hypesthesia
23. skin (194 terms) cafe au lait spots, petechiae, rash
24. sleep (48 terms) cataplexy, narcolepsy, hypersomnia
25. speech language (116 terms) dysarthria, aphasia, echolalia
26. sphincter (67 terms) urinary incontinence, constipation, enuresis
27. tremor (48 terms) tremor, resting tremor, action tremor
28. vision (450 terms) achromatopsia, scotoma, optic atrophy
29. weakness (159 terms) proximal weakness, foot drop, triceps weakness
30. miscellaneous (618 terms) nausea, vomiting, bradycardia

We used python to assign each phenotypic feature (sign or symptom) to one of the thirty superclasses based on the lookup table (see Table 1 for an illustration of how individual phenotype features were mapped to superclasses. The lookup table is available on the project GitHub site.)

### Vectorization (Conversion of phenotype lists to phenotype vectors)

Variable-length lists of phenotypic features were converted into vectors of fixed length 31 elements. The first element of the list was the disease label, and the next 30 elements were the counts of features in each of the 30 superclasses based on the lookup table. When the phenotype is represented as a numerical vector, comparison of phenotypes by various distance metrics is possible. Furthermore, the magnitude of each element in the phenotype vector carries information that facilitates comparisons between diseases. For example, a disease with the hyperkinetic features dystonia, chorea, and athetosis would have a hyperkinesia superclass value of n = 3, whereas a disease with only dystonia would have a hyperkinesia superclass value of n = 1. Such weightings could be useful in distinguishing between phenotypes of similar diseases.

### Visualization (Creation of Heat Maps and Word Clouds)

Heat maps and word clouds were based on the phenotype vectors generated by python. Heat maps were created using the *heat map widget* from Orange [12]. The score mapped for each superclass was the count of the phenotype features subsumed by that class. When a superclass had no features assigned to it, that superclass was dropped from the heat map. Word clouds were produced using the *word cloud widget* from Orange. Word size in the word cloud reflected the frequency of phenotypic features for a group of diseases (Figure 3b) or a single disease (Figure 2b).

**Figure 3b.**
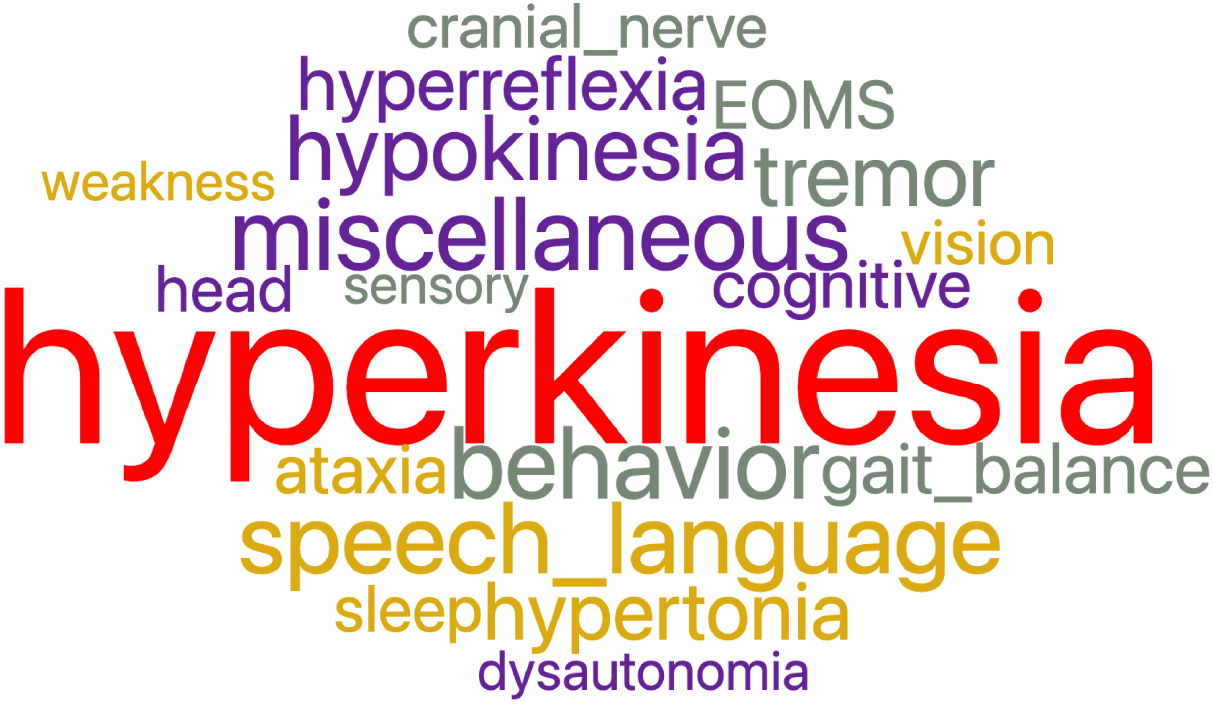
After feature reduction by subsumption, the number of superclasses needed to characterize dystonia diseases was reduced to nineteen. The largest superclass is hyperkinesia which encompasses dystonia, generalized dystonia, focal dystonia, blepharospasm, craniofacial dystonia, and others.

## RESULTS

As our use case, we examined the phenotype profiles of 32 disease variants of dystonia in Orphadata. Phenotype profiles were linear lists of features (see Table 1 for examples of DYT4, DYT6, DYT16, and DYT27). Feature lists ranged between 5 and 48 elements, with a mean of 18.4 features *±* 10.5. The 252 unique features in the phenotype lists were reduced by subsumption into one of 19 superclasses (Table 1 and Figures 3a and 3b). This allowed visualization of the entire dystonia disease set of 32 variants as a heat map (Figure 1). This heat map allows an easy distinction of pure dystonia (e.g., DYT25 and DYT26) from dystonias with sensory loss (e.g., autosomal dominant dopa-responsive dystonia), cognitive impairment (e.g., DYT4) and hypokinesia (e.g., adult-onset dystonia-parkinsonism). Individual rows in the heat map (Figure 2a can be further visualized with word clouds which emphasize phenotypic differences between the dystonia variants (see Figure 2b) for word clouds of DYT4, DY6, DYT16, and DYT 27.

## DISCUSSION

Rich and detailed information on the phenotypes of neurological diseases is held in online repositories such as OMIM, HPO, and Orphadata. Detailed phenotypic data is available for download and can be used to gain insights into the inter-relationships between genes, disease, and phenotypes. Nonetheless, visualization of the phenotypic data retrieved as linear lists remains problematic. We identified limitations to current disease phenotype visualizations, which included:

1. Phenotype feature lists are long.
2. Too many of the phenotype features are near synonyms, hyponyms, or hypernyms.
3. The number of unique features is large.
4. Side-by-side comparisons of phenotype features by diseases are difficult.
5. Phenotype lists of signs and symptoms are co-mingled with radiological, pathological, biochemical, and electrophysiological findings.

To address these limitations, we proposed first restricting our attention to visualizing the phenotypes of rare neurological diseases in Orphadata (N= 1,184). We mapped each of the 4,505 unique features used to describe signs and symptoms in Orphadata into one of 30 superclasses by subsumption (Figures 3a and 3b). This allowed us to convert phenotype lists of variable length to vectors of defined length (31 elements) in which the first element of the vector was the disease label and the next 30 elements were the count of features for each of the 30 superclasses. This process of converting a list to a vector is illustrated in Table 1 for DYT4, DYT6, DYT16, and DYT27. Only 11 of the 30 superclasses were needed to represent these four dystonias. Once phenotype lists are converted to vectors, a group of diseases can be represented as a matrix. For example, 32 dystonic diseases from Orphadata can be converted to a matrix with 32 rows (each row a disease) and 20 columns (each column a superclass of phenotypic features plus one column for the disease label) and then visualized as a heat map (Figure 1). For easy readability, individual rows (diseases) in the heat maps can be converted to word clouds to visualize better the phenotype (Figure 2b).

We have addressed *limitation 1* (long feature lists) by using subsumption to collapse 4,505 phenotypic features into 30 neurological superclasses. This subsumption of numerous phenotypic features into 30 superclasses also addresses *limitation 2* (too many near-synonyms) and *limitation 3* (too many unique features). Once phenotype lists of variable length are converted to vectors of defined length, side-by-side comparisons of diseases become feasible through the use of heat maps and word clouds (Figures 2a and 2b) which addresses *limitation 4*. Another advantage of vectorization is that it allows the calculation of distances between phenotypes using standard distance metrics such as cosine and Euclidean. Figure 1 demonstrates the clustering of rows (dystonic diseases) using the Euclidean distance. We filtered out biochemical, radiological, electrophysiological, and pathological features to address *limitation 5* (excluding features that are not signs and symptoms.)

This work has limitations. Collapsing granular phenotype features into superclasses by subsumption involves information loss. The superclasses retain no laterality information (left-sided versus right-sided weakness, etc.) The superclasses retain no topographical information (proximal versus distal weakness, etc.) The high information value of some granular phenotype features, such as impaired vertical gaze (a sign of progressive supranuclear palsy) or internuclear ophthalmoplegia (a sign of multiple sclerosis), is lost when the granular features are collapsed into the superclass of abnormal eye movements. In addition, our current process of collapsing phenotype concepts into superclasses requires a manually constructed lookup table that assigns each concept to a superclass. Errors can be made in assigning concepts to superclasses. We are looking at ways to improve the subsumption process that collapses ontology concepts into superclasses. Furthermore, the heat map scales are non based on continuous values. For each superclass score, we counted the number of features in that superclass. For example, a disease phenotype with the term *hemiparesis* would have a superclass score of 1 for weakness. In building the features maps, a more general concept like *hyperreflexia* carries the same weight as a more limited concept such as *increased biceps reflex*. In contrast, a disease phenotype with terms *arm weakness* and *leg weakness* would have a superclass score of 2. Moreover, we did not weight phenotype features by importance. We are exploring whether normalization or other data transformations would improve the heat maps.

Another limitation of the work is that the size and granularity of the superclasses are not uniform. For example, the vision superclass subsumed 450 concepts and had many different types of visual impairment, whereas the fatigue superclass subsumed only 26 concepts and reflected the concept of fatigue alone. The selection of the thirty superclasses reflected domain expert opinion and the underlying structure of the ontologies, other useful partitions of the ontology into superclasses are possible. For example, chorea or dystonia could have been distinct superclasses instead of subsumed into hyperkinesia. Speech (e.g., dysarthria) and language disorders (e.g., aphasia) could have been separate superclasses. The superclasses were restricted to neurological terms and neurological diseases. As a result, the heat maps will not be useful in visualizing the phenotypes of non-neurological diseases.

Furthermore, the heat maps will not adequately visualize important non-neurological signs and symptoms of diagnostic value (such as Kayser-Fleisher rings for Wilson’s disease [15]). Although true pathognomonic signs and symptoms are rare in neurology [26, 27, 3, 9], the heat maps lack the granularity to show pathognomonic signs. The current heat maps do not support a drill down to the underlying granular phenotype features. Although we used Orange to create the heat maps, suitable heat maps are also available in python, and R. Other heat map color schemes are available and may have given better visualizations. The Orphadata phenotype datasets are undergoing revisions and improvements. Some diseases are phenotyped more completely than others. Although the dataset is curated, omissions, errors, and discrepancies can still occur. Finally, a similar analysis could have been done with phenotypic annotations from the OMIM or HPO datasets.

## CONCLUSIONS

Combining feature reduction by subsumption with vectorization of phenotype lists and the visualization by heat maps and word clouds offers a robust method to explore neurology phenotypes. Subsumption permits the reduction of thousands of ontological concepts into a reduced number of phenotype superclasses. Vectorization allows the conversion of phenotype feature lists of variable length into superclass vectors of defined length. Matrices of superclass vectors allow the side-by-side comparison of neurological disease phenotypes as heat maps. Entire heat maps or individual rows can be visualized as word clouds, providing an easy-to-grasp representation of neurological disease phenotypes.

## Data Availability

All data produced in the present study are available upon reasonable request to the authors.

https://github.com/dbhier/visualization_of_neurology_phenotypes

## AUTHOR CONTRIBUTIONS

Concept by DBH and RY. Data analysis by DBH and RY. Data interpretation by DBH, MDC, RY, and DCW III. Writing, revision, and approval by DBH, MDC, RY, and DCW III.

## ACKNOWLEDGMENTS

We acknowledge helpful discussions with our colleagues Chelsea Oommen, Quentin Howlett-Prieto, and Fahime Shojaei.

## DATA AVAILABILITY STATEMENT

Underlying data is available at the project’s GitHub site at github.com/dbhier/visualization of neurology phenotypes

